# Trio-based GWAS identifies novel associations and subtype-specific risk factors for cleft palate

**DOI:** 10.1101/2023.03.01.23286642

**Authors:** Kelsey Robinson, Trenell J. Mosley, Kenneth S. Rivera-González, Christopher R. Jabbarpour, Sarah W. Curtis, Wasiu Lanre Adeyemo, Terri H. Beaty, Azeez Butali, Carmen J. Buxó, David J. Cutler, Michael P. Epstein, Lord JJ Gowans, Jacqueline T. Hecht, Jeffrey C. Murray, Gary M. Shaw, Lina Moreno Uribe, Seth M. Weinberg, Harrison Brand, Mary L. Marazita, Robert J. Lipinski, Elizabeth J. Leslie

## Abstract

Orofacial clefts (OFCs) are the most common craniofacial birth defects and are often categorized into two etiologically distinct groups: cleft lip with or without cleft palate (CL/P) and isolated cleft palate (CP). CP is highly heritable, but there are still relatively few established genetic risk factors associated with its occurrence compared to CL/P. Historically, CP has been studied as a single phenotype despite manifesting across a spectrum of defects involving the hard and/or soft palate. We performed GWAS using transmission disequilibrium tests using 435 case-parent trios to evaluate broad risks for any cleft palate (ACP, n=435), as well as subtype-specific risks for any cleft soft palate (CSP, n=259) and any cleft hard palate (CHP, n=125). We identified a single genome-wide significant locus at 9q33.3 (lead SNP rs7035976, p=4.24×10^−8^) associated with CHP. One gene at this locus, angiopoietin-like 2 (*ANGPTL2*), plays a role in osteoblast differentiation. It is expressed in craniofacial tissue of human embryos, as well as in the developing mouse palatal shelves. We found 19 additional loci reaching suggestive significance (p<5×10^−6^), of which only one overlapped between groups (chromosome 17q24.2, ACP and CSP). Odds ratios (ORs) for each of the 20 loci were most similar across all three groups for SNPs associated with the ACP group, but more distinct when comparing SNPs associated with either the CSP or CHP groups. We also found nominal evidence of replication (p<0.05) for 22 SNPs previously associated with cleft palate (including CL/P). Interestingly, most SNPs associated with CL/P cases were found to convey the opposite effect in those replicated in our dataset for CP only. Ours is the first study to evaluate CP risks in the context of its subtypes and we provide newly reported associations affecting the broad risk for CP as well as evidence of subtype-specific risks.

## Introduction

Orofacial clefts (OFCs) are the most common craniofacial congenital anomalies in humans, occurring at a birth prevalence of ∼1 in 1000 live births(1, 2). Prognosis is favorable with surgical intervention, though individuals with OFCs face many healthcare challenges such as multiple corrective surgeries, abnormal dentition, hearing and speech problems, and increased morbidity and mortality throughout life(1, 3). As such, these anomalies are associated with increased healthcare costs and long-term psychosocial burdens for individuals and their families(4).

OFCs are typically classified into groups including cleft lip (CL), cleft lip with cleft palate (CLP), and cleft palate (CP). Etiologically, cleft lip with or without a cleft palate (CL/P) is considered anatomically and epidemiologically distinct from CP, although both are highly heritable(5, 6) with estimates up to 90% for both CL/P and CP(7). However, compared to dozens of known risk loci for CL/P(8, 9), common risk variants for CP remain largely undiscovered. Large-scale studies of CP, including seven genome-wide association studies (GWAS)(10-16), have mostly identified variants occurring either at low frequency or in specific populations. For example, a missense variant in *GRHL3* occurs at ∼3% only in Europeans(10), a variant near *CTNNA2* occurs at ∼1.5% in an African population(14), and a variant in an enhancer region of *IRF6* occurs almost exclusively in Finnish and Estonian Europeans(16).

Historically, CP has been evaluated as a single group, but it encompasses a phenotypic spectrum including overt clefts of the hard and/or soft palate and submucous cleft palate. There is evidence for specific gene expression patterns in the hard versus soft palate(17), so we hypothesized there may be underlying genetic differences among CP subtypes. To investigate this further, we performed GWAS of three groups of case-parent trios via transmission disequilibrium tests (TDT). These groups include all cases of CP regardless of subtype, cases involving the soft palate (i.e., hard and soft palate, soft palate only, and submucous cleft palate), and cases involving the hard palate (i.e., hard and soft palate plus hard palate only).

## Results

### GWAS of Any CP Type

We performed TDT using all 435 CP case-parent trios (Supplemental Table 1) for 6,946,419 variants (Supplemental Figure 1). In the combined analysis of all trios with CP (hereafter referred to as any CP, or ACP), no loci reached genome-wide significance (p<5×10^−8^), although there were 10 loci with at least one SNP surpassing a suggestive threshold of p<5×10^−6^ (Table 1). Several GWAS of CP have been published to date (10-16), however we did not find any of these previous loci beyond nominal significance in this study, as further discussed below (Supplemental Table 2).

**Table 1.**
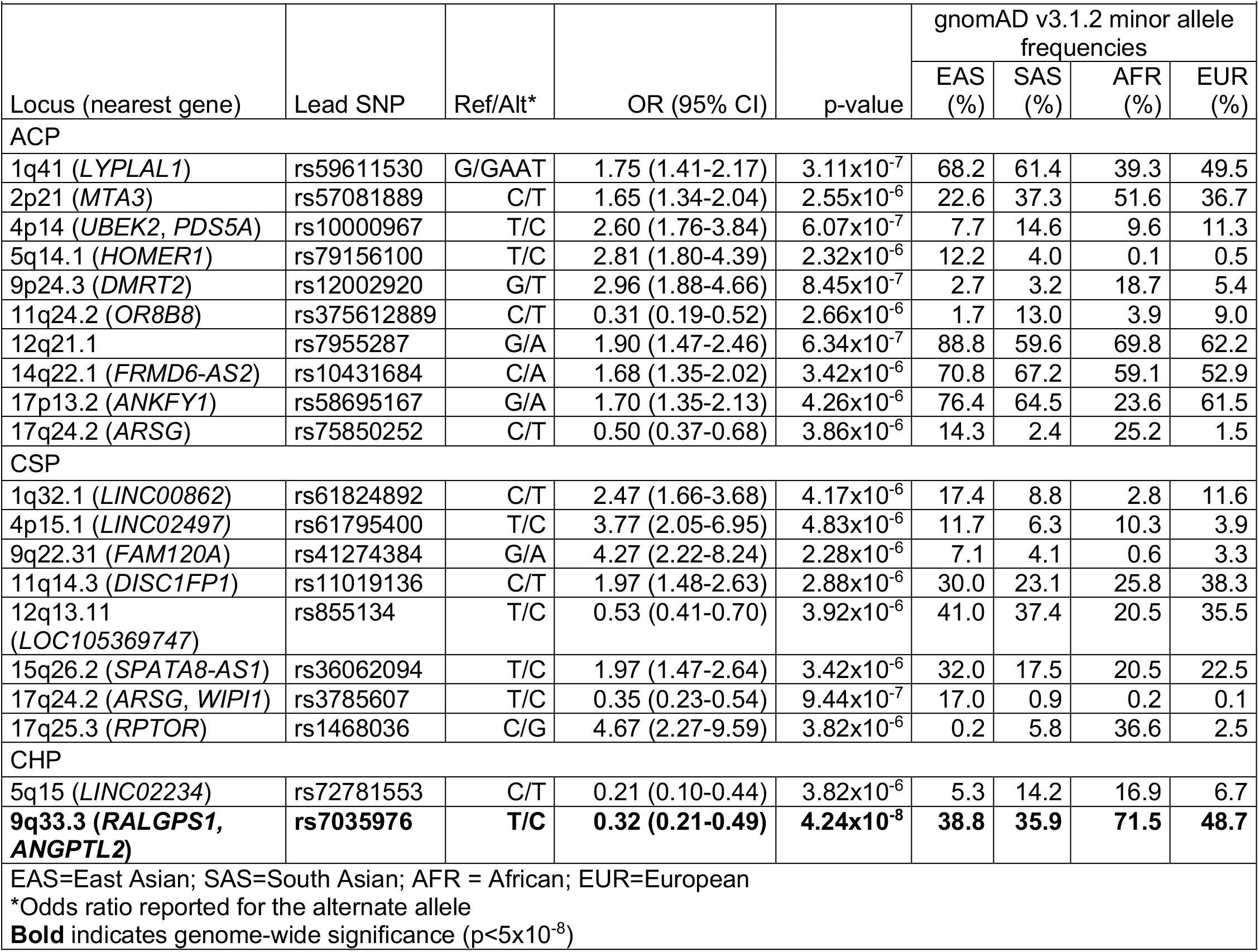
Suggestive and significant loci from any CP type and subtype-specific GWAS.

### Subtype-Specific GWAS

CP is phenotypically heterogenous, where clefts can occur in the hard palate (the bony, anterior portion) and/or the soft palate (the muscular, posterior portion). Given that such phenotypic distinctions arise from different developmental origins, we hypothesized some of the relative lack of associated SNP signals could be attributed to this underlying heterogeneity. To determine if CP subtypes were associated with unique loci, we performed TDTs on two subgroups: “any cleft of the soft palate” which included 259 trios with clefts of the hard and soft palate, soft palate only, or submucous cleft (CSP) (Supplemental Figure 2) and “any cleft of the hard palate” which included 125 trios with hard and soft palate or hard palate only (CHP) (Supplemental Figure 3). Those with clefts of both the hard and soft palate were included in both analyses because that group may be etiologically heterogeneous and may have hard palate and/or soft palate risk factors.

The CSP analysis included 7,286,217 variants. There were no loci reaching genome-wide significance, although there were 8 loci of suggestive significance (Table 1). Our CHP analysis included 7,337,001 variants, and we identified a genome-wide locus on chromosome 9q33.3 (Figure 1A, 1B) spanning the genes *RALGPS1* and *ANGPTL2*, as well as one locus of suggestive significance (Table 1). We used FINEMAP (18) to perform statistical fine-mapping of the 9q33.3 locus, which identified three SNPs within the credible set for which there was 100% confidence at least one is association with disease: rs2417050, rs777676, rs12350252 (Figure 1C).

**Figure 1:**
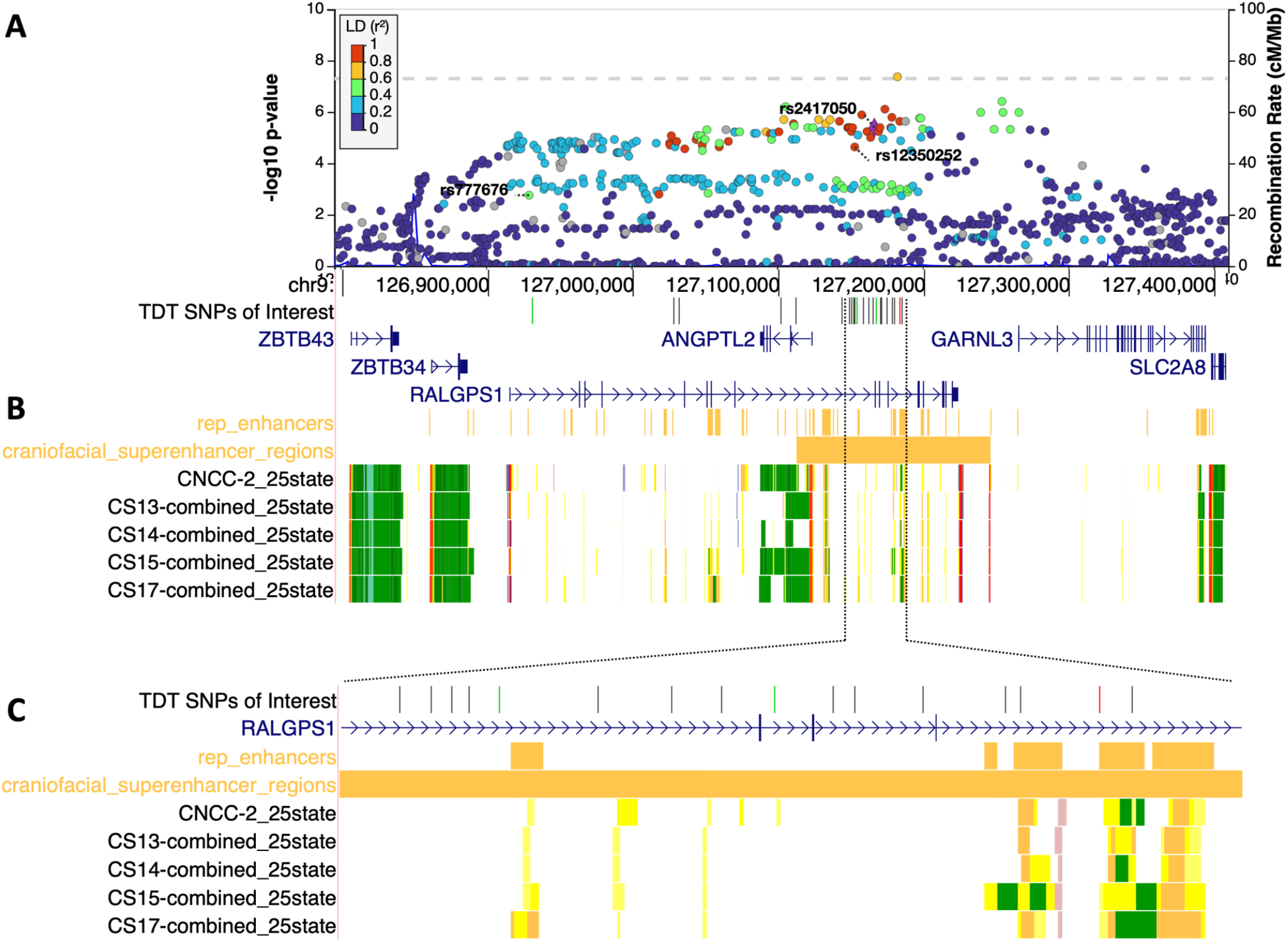
Genome-wide significant locus at 9q33.3 spans craniofacially-expressed gene *ANGPTL2* and a craniofacial superenhancer. A) Regional association plot for 9q33.3. The labelled SNPs were identified by FINEMAP with 100% confidence of belonging to the credible set of SNPs associated with disease. B) UCSC genome browser tracks (http://genome.ucsc.edu) for craniofacial-specific gene expression and regulatory regions. C) Zoomed in view of the region with high density of SNPs in LD with SNPs labelled in 1A. Point color corresponds to linkage disequilibrium (r^2^) with rs2417050 across all populations. For browser tracks, yellow indicates an enhancer region (darker shades represent stronger elements), green indicates active transcription, and red indicates a transcription start sites. CNCC=cranial neural crest cells, CS=Carnegie stage.

We next compared the CSP and CHP analyses to each other and to the ACP group to determine to what extent these loci were associated with a specific CP subtype. The only overlap in suggestive loci was 17q24.2 (near *ARSG*), shared by the ACP (lead SNP rs75850252, 2.89×10^−6^) and CSP analyses (lead SNP rs3785607, p=9.44×10^−7^). The 17q24.2 signal was driven by the CSP group but also showed nominal evidence of association in CHP (p=0.006). Due to the overlap in samples, however, we cannot distinguish between an association of this locus with any type of CP or with a cleft of both the hard and soft palate. There were additional regions, such as the 1q41 locus shown in Figure 2A, in which the association patterns were similar between ACP, CSP, and CHP, but did not reach the suggestive threshold in the subtype analyses. In contrast, we observed loci with stronger association signals in one subtype versus the other or in ACP. For example, the top locus in CHP, 9q33.3, reached genome-wide significance but this signal was less significant in CSP (p=2.28×10^−4^) and ACP (p=1.79×10^−3^) (Figure 2B), suggesting this signal was driven by clefts of the hard palate (either with or without cleft soft palate). Similarly, the 9q22.31 locus identified in CSP is less significant in ACP (p=5.41×10^−4^) and was essentially absent when looking at CHP (p=0.53) (Figure 2C), suggesting it is driven exclusively by cleft soft palate.

**Figure 2:**
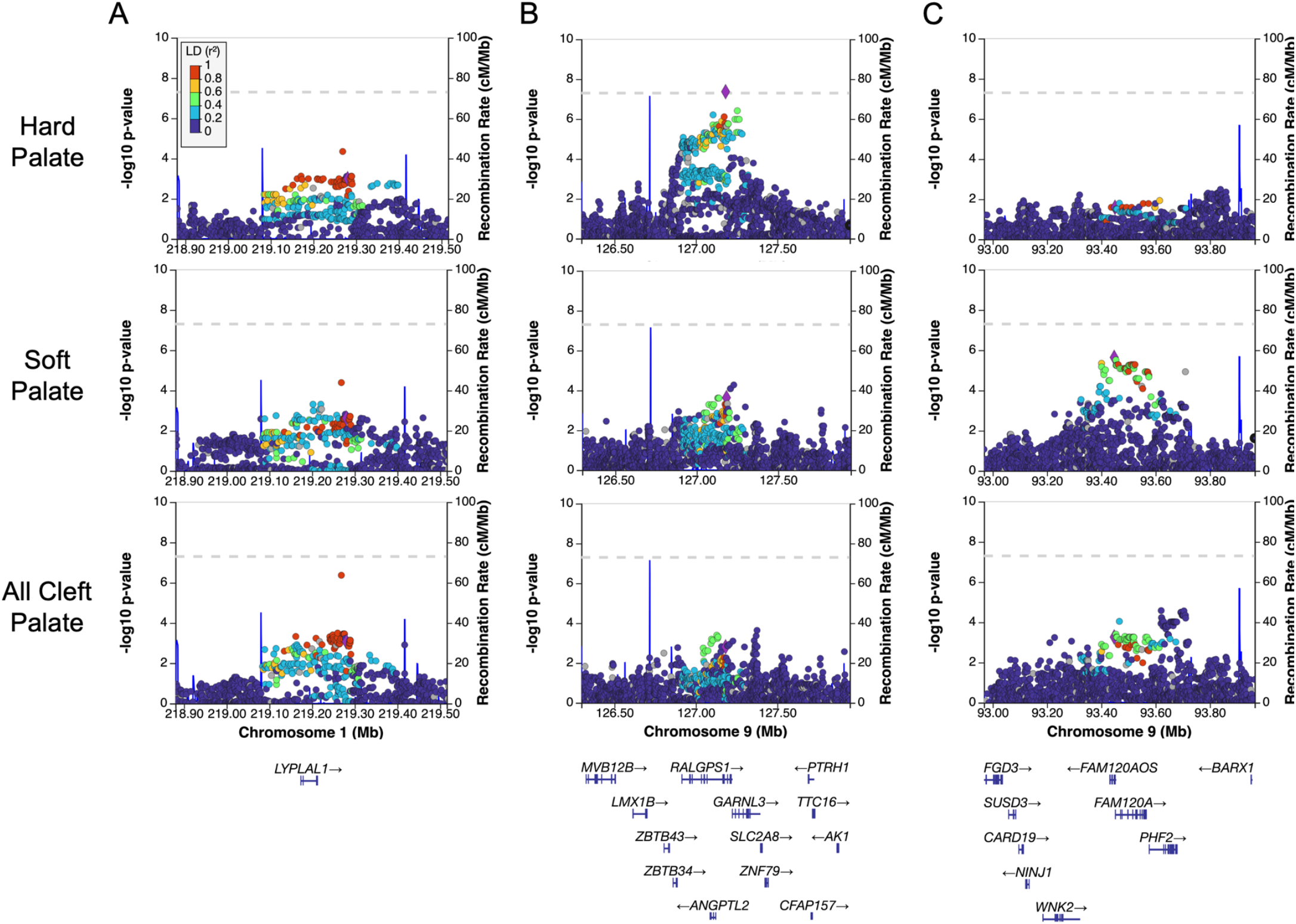
Regional association plots illustrate differences between groups at A) the 1q41 locus spanning *LYPAL1* with index SNP rs10779347 B) the 9p33.3 locus spanning *RALGPS1* with index SNP rs7035976, and C) the 9q22.31 locus spanning *FAM120A* with index SNP 4127438 demonstrating similar association patterns (A) and subtype specific associations (B, C). Point color corresponds to linkage disequilibrium (r^2^) and the blue lines represent linkage block boundaries.

We then compared the estimated odds ratios (ORs) for the lead SNPs in each region showing suggestive significance for the three subgroups (Figure 3) to identify subtype-specific risks suggested by comparisons of p-values. As predicted, the ORs were very similar across all groups for loci identified in the ACP analysis. However, for loci identified in either CSP or CHP, the differences in ORs were more pronounced. Although most confidence intervals overlap between CSP and CHP, there was some evidence for subtype-specific effects. For the SNPs identified from the CHP analysis, there was no overlap in the range of effects for the 5q15 locus in CHP and CSP (which also contained 1), indicating this locus is specific to clefts involving the hard palate. It is less clear for the SNPs identified in the CSP analysis. For 4 of the 10 SNPs, the confidence interval for CHP contained 1, which allows the possibility of no effect in that group. Additionally, all of the confidence intervals overlapped, indicating these loci may not be subtype-specific. However, taken altogether, these findings suggest there are subtype-specific genetic risks for CP.

**Figure 3:**
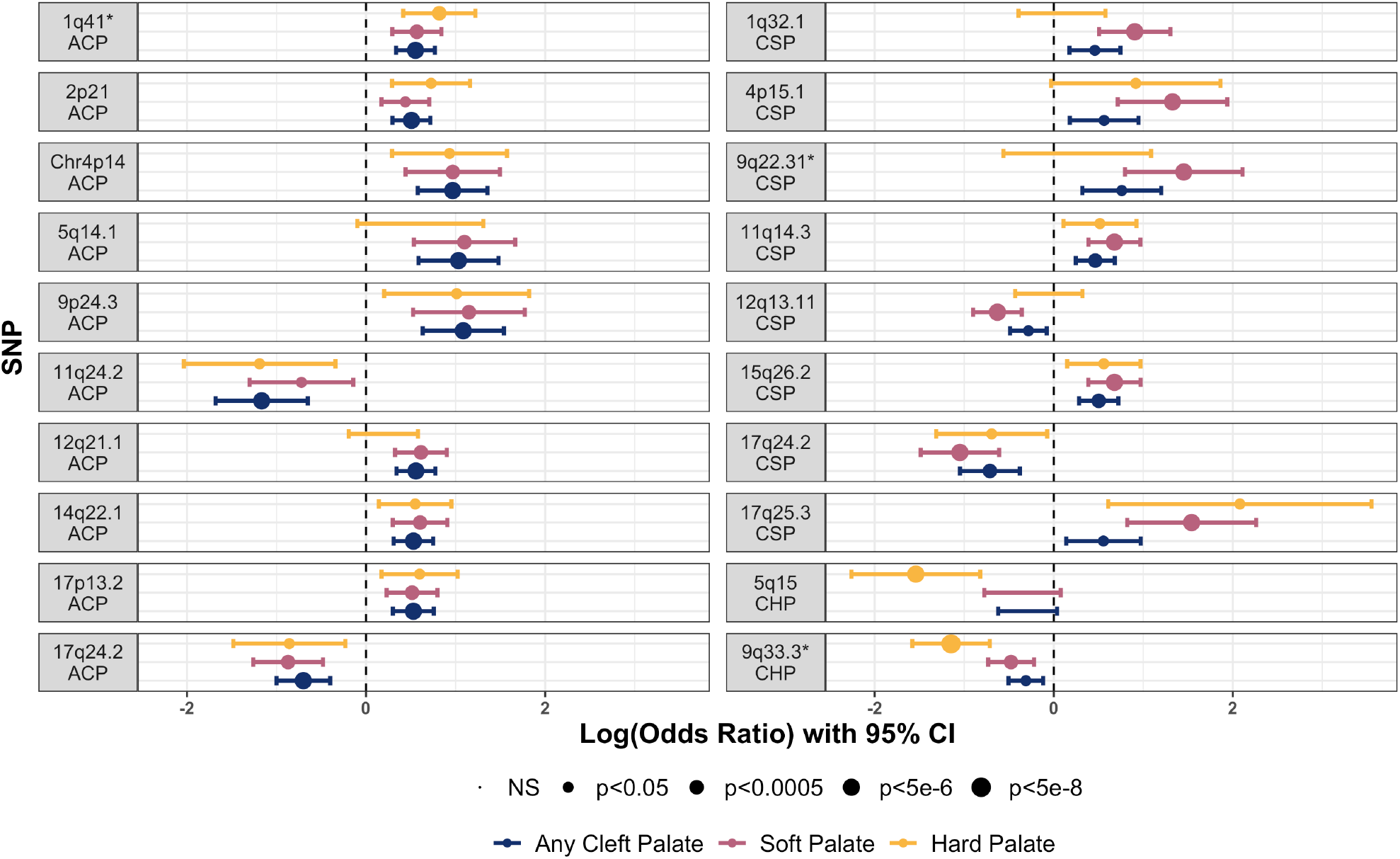
Comparison of odds ratios for any suggestive loci demonstrates subtype-specific effects. Loci associated with any cleft palate convey similar ORs for both subtypes (left). Loci associated with a specific subtype (right) carry less extreme ORs and/or are insignificant in the opposing group. Loci marked with an asterisk are featured in Figure 2. ACP=any cleft palate. CSP=cleft soft palate. CHP=cleft hard palate.

Lastly, because our cohort represents diverse populations, we evaluated population-specific signals. Although our test of homogeneity did not show any significant differences between each analysis (p=0.137), differences in site heterozygosity between ancestries can mask signals. Using principal component analysis, we subdivided groups into Asian, European, and African ancestry groups (Supplemental Figure 4); however, only the Asian subgroup contained enough trios (N=262) to perform TDT for 6,491,466 variants (Supplemental Figure 5). The results from this analysis did not reveal any additional loci not already present in the full cohort, but did demonstrate that some signals in the full cohort, such as 5q14.1 are likely driven by this population. This is also supported by the population frequencies in Table 1.

### Expression during mouse palatogenesis and limb development

We further investigated the 9q33.3 locus, as the only genome-wide significant locus, to locate a candidate gene for cleft palate. Based on available expression and chromatin segmentation data from human embryonic craniofacial tissue (19), *ANGPTL2* is actively transcribed in neural crest cells and human craniofacial tissue during embryogenesis at Carnegie Stages 13, 14, 15, and 17, whereas this pattern is not apparent for *RALGPS1* (Figure 1B). We therefore hypothesized *ANGPTL2* was more likely than *RALGPS1* to be involved in palatal development (Figure 1B). We performed *in situ* hybridization on mouse embryos at two key stages of palatogenesis: initial vertical outgrowth of the palatal shelves, and subsequent horizontal outgrowth just prior to their fusion (20). *Angptl2* staining was observed in the palatal shelves and the lateral aspects of the upper lip at both stages of development (Figure 4A, 4B). Tissue sectioning revealed *Angptl2* staining in the mesenchymal compartment of the palatal shelves (Figure 4A’, 4B’). Subsequent quantitative assessment demonstrated that mesenchymal *Angptl2* expression increases during palatogenesis in a manner consistent with that of *Runx2*, an established marker of osteogenic differentiation (Figure 4C); this is consistent with our observation that the association was driven by clefts of the hard palate.

**Figure 4.**
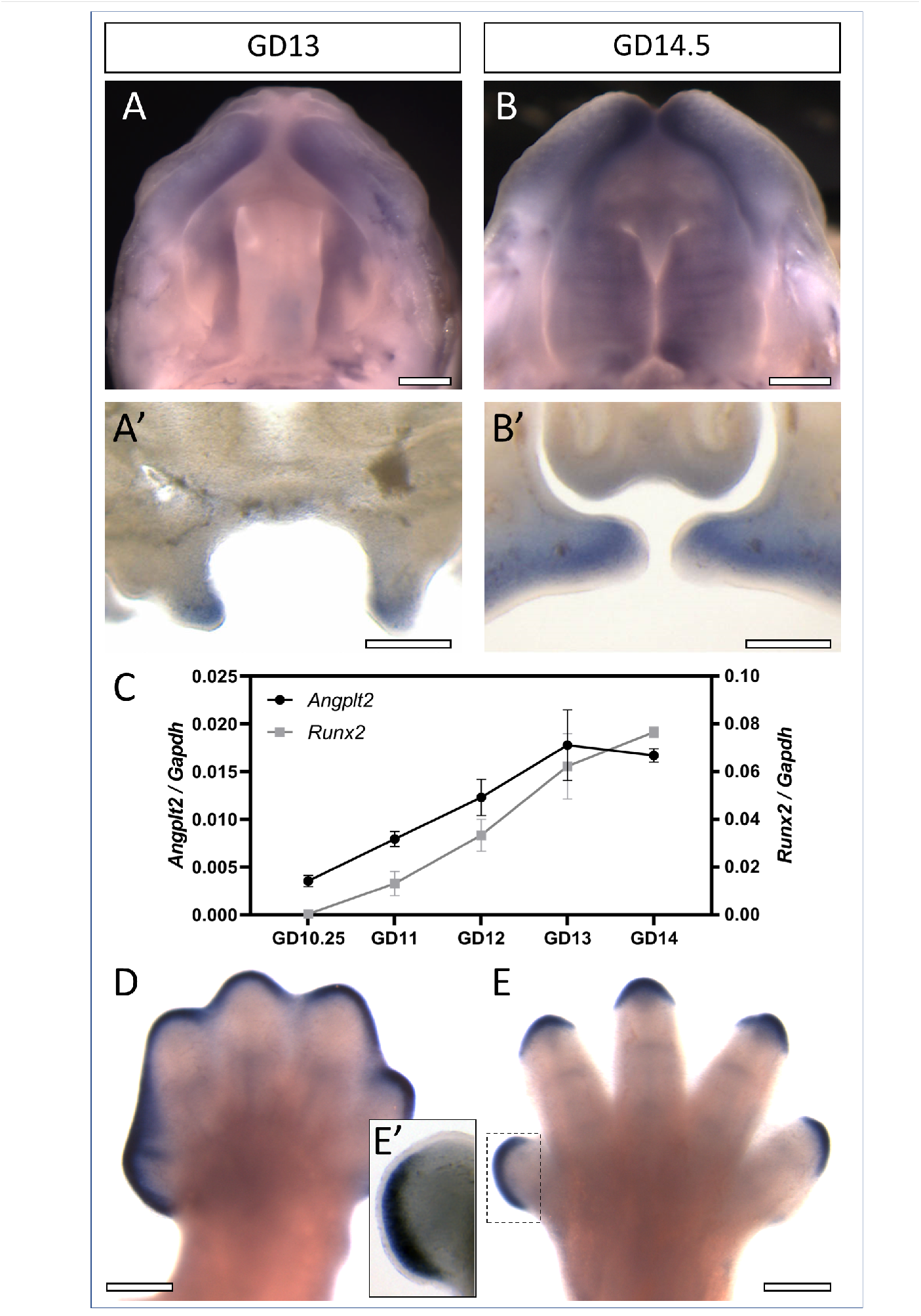
*Angptl2* expression during mouse palate and limb development. Mouse embryos at gestational day (GD)13 or 14.5 were stained by *in situ* hybridization to visualize *Angptl2* expression. Whole mount tissues were imaged to view the developing palatal shelves and upper lip (A,B). Subsequent coronal sections illustrate prominent staining in mesenchymal tissue of the palatal shelves (A’,B’). qPCR was conducted on mesenchyme isolated from maxillary process/palatal shelf tissue from mice at indicated time points (C). Each value represents the mean ± SEM of n=3 samples isolated from individual embryos. Forelimbs from GD13 and 14.5 mouse embryos were also stained by *in situ* hybridization to visualize *Angptl2* expression (D,E). A section through the first digit illustrates staining restricted to the mesenchyme adjacent to apical ectodermal ridge. Similar domains of expression were observed in hindlimbs. Scale bars = 0.50 mm.

We also observed strong *Angptl2* staining during limb development in a domain restricted to the mesenchyme adjacent to the apical ectodermal ridge (Figure 4D, E). The apical ectodermal ridge secretes signals that maintain the adjacent mesenchyme (i.e., progress zone) in a highly proliferative state, driving proximodistal outgrowth of the limb and digits (21). We therefore investigated overlapping phenotypes using the DECIPHER database (22). We found 38 reported individuals with copy number variants (CNVs) affecting *ANGPTL2* and adjacent genes in this region. Of these, 13/38 (34.2%) had craniofacial phenotypes (e.g., cleft palate, narrow palate, high palate) and 15/38 (39.5%) had limb abnormalities (e.g., camptodactyly of finger, arachnodactyly) with an overlap of 10/38 (26.3%) with clinical features of both craniofacial and limb abnormalities. Using prevalence data from EUROCAT, we compared the rate of OFCs (14.95 per 10,000 births) and limb anomalies (38.18 per 10,000 births) in the general population to that of patients with CNVs of *ANGPTL2* and found a significant difference between these two groups (p<2.2×10^−16^, Fisher’s exact two-tailed test) for both OFCs and limb anomalies.

### Attempted replication of previous published SNPs

We investigated SNPs from previously published studies of OFCs for evidence of replication in our dataset. Using a list from the GWAS Catalog (23), we tested 139 unique SNPs for association with ACP, CSP, or CHP. There were 22 SNPs within 18 loci achieving nominal significance (p<0.05) in at least one group from our analyses (Table 2, Supplemental Table 2). When we applied Bonferroni multiple-test correction (p<3.59×10^−4^), a single SNP (rs1838105, p=2.95×10^−4^) remained significant for replication in the ACP group.

**Table 2:** Previous published OFC-associated SNPs with evidence of replication in the current study.

|  |  |  |  |  |  |  |  |  |  |  |  |  |
| --- | --- | --- | --- | --- | --- | --- | --- | --- | --- | --- | --- | --- |
| Previous Study |  |  |  |  |  |  | Current Study |  |  |  |  |  |
| Variant | Locus (hg38) | Alleles | Effect Allele* | OR (95% CI) | Trait | PubMed ID | CP OR (95% CI) | CP p-value | CSP OR (95% CI) | CSP p-value | CHP OR (95% CI) | CHP p-value |
| rs12065278 | 1p36.11 | A/G | G | 2.43 (1.66-3.56) | CP | 27018472 | 1.18 (0.92-1.52) | 2.00E-01 | 1.22 (0.89-1.67) | 2.25E-01 | 1.63 (1.04-2.57) | **3.25E-02** |
| rs481931 | 1p22.1 | G/T | G | 1.25 | CLP | 28232668 | 1.25 (1.01-1.54) | **3.96E-02** | 1.22 (0.93-1.61) | 1.61E-01 | 1.08 (0.70-1.61) | 7.51E-01 |
| rs4839542 | 1p13.3 | C/T | C | 0.83 | CLP | 28232668 | 1.23 (0.95-1.59) | 1.14E-01 | 1.22 (0.88-1.72) | 2.33E-01 | 1.96 (1.20-3.12) | **5.27E-03** |
| rs2235371 | 1q32.2 | C/T | T | 1.49 (1.37-1.61) | CL/P | 25775280 | NA | NA | 1.38 (1.0-1.89) | **4.60E-02** | 1.53 (1.0-2.33) | **4.64E-02** |
| rs6540559 | 1q32.2 | G/A | A | 1.67 (1.47-1.85) | CL/P | 28054174 | 1.24 (1.01-1.52) | **3.57E-02** | 1.30 (1.0-1.70) | 5.11E-02 | 1.48 (0.99-2.2) | 5.62E-02 |
| rs75477785 | 1q32.2 | T/G | G | 1.75 (1.54-2.04) | CL/P | 28054174 | NA | NA | 1.38 (1.0-1.9) | 5.16E-02 | 1.61 (1.04-2.48) | **3.10E-02** |
| rs72741048 | 1q32.2 | A/T | T | 1.31 | CP | 31609978 | 1.35 (1.09-1.66) | **5.58E-03** | 1.28 (0.97-1.67) | 7.55E-02 | 1.67 (1.15-2.44) | **6.85E-03** |
| rs11119394 | 1q32.2 | A/G | G | 1.30 | CP | 32758111 | NA | NA | 1.48 (1.09-2.01) | **1.16E-02** | 1.46 (0.98-2.18) | 5.87E-02 |
| rs3815854 | 2q35 | C/T | C | 0.80 (0.74-0.87) | CL/P | 25775280 | 1.25 (1.03-1.52) | **2.45E-02** | 1.35 (1.06-1.72) | **1.38E-02** | 1.19 (0.83-1.72) | 3.53E-01 |
| rs9347594 | 6q26 | T/C | T | 2.22 (1.60-3.08) | CP | 27018472 | NA | NA | 1.11 (0.83-1.47) | 4.77E-01 | 1.49 (1.0-2.22) | **4.77E-02** |
| rs11774066 | 8q22.2 | C/T | C | 0.86 | CLP | 28232668 | 1.28 (1.03-1.61) | **2.44E-02** | 1.16 (0.87-1.54) | 3.07E-01 | 1.09 (0.741.59) | 6.92E-01 |
| rs1487022 | 8q22.2 | G/T | G | 1.17 | CLP | 28232668 | 1.45 (1.16-1.85) | **1.36E-03** | 1.47 (1.08-2.0) | **1.58E-02** | 1.14 (0.76-1.72) | 5.27E-01 |
| rs4246129 | 8q24.3 | C/G | G | 2 (1.47-2.63) | CP | 28054174 | 1.28 (1.03-1.61) | **2.75E-02** | 1.47 (1.1-1.97) | **8.65E-03** | 1.16 (0.77-1.74) | 4.73E-01 |
| rs7928246 | 11q22.3 | A/G | A | 0.41 | CP | 32758111 | 1.77 (1.05-2.99) | **2.95E-02** | 2.18 (1.07-4.45) | **2.80E-02** | NA | NA |
| rs730643 | 14q13.3 | G/A | A | 1.35 | CP | 31609978 | 1.34 (1.09-1.66) | **5.85E-03** | 1.38 (1.05-1.82) | **2.18E-02** | 1.60 (1.05-2.44) | **2.77E-02** |
| rs4901118 | 14q22.1 | G/A | G** | NR | CL/P | 28087736 | 1.41 (1.16-1.72) | **4.57E-04** | 1.45 (1.12-1.89) | **4.04E-03** | 1.19 (0.84-1.69) | 3.29E-01 |
| rs152745 | 16p12.2 | G/A | G | 0.46 (0.33-0.64) | CP | 27018472 | NA | NA | 1.19 (0.88-1.59) | 2.57E-01 | 1.61 (1.04-2.50) | **2.91E-02** |
| rs57933945 | 16p12.1 | C/T | C | 2.86 (1.85-4.35) | CP | 28054174 | 1.92 (1.32-2.86) | **6.25E-04** | 2.5 (1.45-4.35) | **6.70E-04** | 2.22 (1.04-4.55) | **3.39E-02** |
| rs9911652 | 17p13.1 | C/T | C | 0.64 (0.55-0.74) | CL/P | 28054174 | 1.33 (1.01-1.75) | **3.94E-02** | 1.3 (0.92-1.85) | 1.35E-01 | 1.37 (0.85-2.17) | 1.92E-01 |
| rs3785888 | 17q21.32 | C/T | T | 0.88 (0.58-0.97) | CL/P | 28054174 | 1.28 (1.06-1.55) | **9.13E-03** | 1.19 (0.93-1.51) | 1.59E-01 | 1.08 (0.76-1.53) | 6.60E-01 |
| rs1838105 | 17q21.32 | A/G | G | 0.82 | CLP | 28232668 | 1.44 (1.18-1.75) | **2.40E-04** | 1.34 (1.05-1.72) | **2.00E-02** | 1.32 (0.9-1.93) | 1.51E-01 |
| rs227731 | 17q22 | T/G | <u>G</u> | 0.81 (0.72-0.89) | CL/P | 28054174 | 1.30 (1.06-1.59) | **1.07E-02** | 1.27 (0.97-1.64) | 8.24E-02 | 1.59 (1.08-2.33) | **1.77E-02** |
| *Effect allele is reported for CPSeq data: underlines indicate opposite effect reported in original publication.<br>**Effect allele for original publication is not reported.<br>**Bold** indicates $p < 0.05$<br>NA indicates absence of data at this SNP location in the CPSeq data.<br>NR indicates absence of data from original publication. | | | | | | | | | | | | |

## Discussion

CP has historically been evaluated as a single phenotype, but here we have identified CP subtype-specific risk factors, including one genome-wide significant locus and 19 regions of suggestive significance. Our genome-wide significant locus—found in the CHP group—spans both *RALGPS1* and *ANGPTL2* genes on 9q33.3. Although fine-mapping analysis did not implicate any single gene as all three of the SNPs in the credible set fall within intronic regions of *RALGPS1*; two SNPs (rs2417050 and rs12350252), as well as the lead SNP at this locus, also fall within a region considered a craniofacial super enhancer upstream of *ANGPTL2*. We then showed using *in situ* hybridization that *Angptl2* was expressed in the developing mouse palate.

Specifically, we found that *Angptl2* is expressed during initial vertical and subsequent horizontal outgrowth of the palatal shelves (Figure 4A, 4B) prior to the approximation and fusion at the midline that forms the secondary palate. Expression appeared restricted to the mesenchymal compartment, which is primarily comprised of cranial neural crest cells that rapidly proliferate to drive palatal shelf outgrowth, and differentiate into osteoblasts that form the bones of the hard palate. As palatogenesis proceeded, mesenchymal *Angptl2* expression increased along with osteoblast marker *Runx2* (Figure 4C), consistent with previous evidence suggesting that *ANGPTL2* positively regulates osteoblast differentiation (25). These data are also consistent with bulk RNAseq from human craniofacial tissue at CS 13, 14, 15, and 17 showing a similar increase with time, and single-cell RNAseq of the mouse palate showing *Angptl2* is expressed in cells consistent with osteoprogenitors at GD15.5 (26). These findings, in combination with significantly higher rates of OFCs with CNVs of this locus, support a role for *ANGPTL2* in palatal development, particularly as related to the hard palate.

The most strongly associated locus in ACP was located in an intergenic region at 12q21.1, closest to the gene *TRHDE* (∼330kb upstream). The second most significant locus was on chromosome 4p14. Although also in an intergenic region, the nearest genes are *UBEK2* and *PDS5A* (∼20kb upstream and downstream, respectively), both of which are strongly expressed in craniofacial tissue during embryogenesis (19). However, *PDS5A*, which plays a role in sister chromatid cohesion during mitosis, is of particular interest: both null and heterozygous loss in mice leads to a cleft palate phenotype with variable expressivity and penetrance (27).

In the CSP group, the top non-overlapping locus was at 9q22.31 spanning *FAM120A*, and RNA-binding protein (28). Although its function has been primarily studied in the context of gastric carcinoma, *FAM120A* plays a role in protecting against oxidative stress-induced apoptosis. Additionally, it has been shown to directly bind insulin-like growth factor II (*IGF-II*) mRNA (29), which is spatially and temporally expressed in developing murine secondary palate (30), and can result in cleft palate—among other clinical features—when dysregulated (31). *FAM120A* is expressed in both mouse soft palate tissue (26) and in human craniofacial tissue during embryological development (19).

Another locus of interest from this analysis is at 12q13.11, in which the lead SNP is approximately 650kb away from *COL2A1*. Variants within this gene are well established as causal for Stickler syndrome, in which CP is a common feature (32); however, our associated signal does not appear to be within the same topological associated domain (TAD) as *COL2A1*, at least in embryonic stem cells, so additional evaluation of this finding is needed.

Because differences in sample sizes (and statistical power) prevent direct comparisons of p-values, we compared odds ratios between analysis groups. We found that for all loci, the risks between the ACP, CHP, and CSP groups were either in the same direction or null (i.e., no loci conveyed opposite effects for different groups). For loci belonging to the ACP group, the ORs and confidence intervals were similar for CHP and CSP; however, for loci identified in the CSP or CHP analyses, differences in ORs were more pronounced. For example, the 5q14.1 region was associated solely with CHP. Differences between estimated ORs of the CSP loci were less apparent, although 4 of the 10 SNPs may have no risk in CHP. Because so many individuals in the dataset had a cleft hard and soft palate and were included in both the CHP and CSP analyses, the overlapping confidence intervals were expected. We cannot rule out influence of CSP SNPs on CHP risk because cleft hard palate only is not as common as other forms of CP and much larger sample sizes will be required to contrast cleft hard palate only and cleft soft palate only. Overall, our results suggest some variants could contribute more to risk to a specific type of CP.

Although the allelic TDT is not confounded by population stratification, examination of allele frequencies across populations suggests some of our findings may be, in part, driven by certain populations. The most pronounced of these occurs for rs1468036 (effect allele G) at 17q25.3, which is found at a frequency of 36.6% in African populations, and less than 6% in other populations studied here. Another example is the 5q14.1 locus near *HOMER1*, which reached suggestive significance in the ACP group (p=2.32×10^−6^) but is more significant in the Asian ancestry-stratified study (p=8.35×10^−7^) and occurred with a minor allele frequency of 12.2% in East Asian populations compared to 4% in South Asians and <0.5% in the remaining study populations. Presently, ancestry-specific analysis was only possible for the Asian population, but future studies on both additional ancestral risks as well as combined stratification by ancestry and subtype would be of interest; however, limited sample sizes in this study preclude these evaluations here.

All of the risk loci identified in this study were of novel association with CP, although three of these of these loci have been previously reported in studies associated with CL/P. Both 9q22.31 and 5p14.1 have been reported as suggestive in a consanguineous GWAS of 40 families (33), and the 12q21.1 region was reported in a Chinese Han population (12); however, the lead SNPs for each of these loci are approximately 1Mb away from each of our lead SNPs. Additionally, none of the identified SNPs within the same region are in linkage disequilibrium, and therefore may or may not be tagging the same causal variant(s).

We were able to show nominal evidence of replication for 22 previously published SNPs associated with OFCs. Interestingly, there were only 2 SNPs from previous studies that replicated in all three of our studies (i.e., p<0.05 in ACP, CSP, and CHP), both of which were originally published as associated with CP. There were 9 previously published SNPs replicating at nominal significance in any two of our groups, only one of which was shared between only CSP and CHP. Unfortunately, deeper phenotype data is not available to classify subtypes from previous studies for more detailed comparisons, but this general lack of overlap may further support subtype-specific differences. An additional striking finding was that for all 7 previously published SNPs with reported ORs associated with CL/P, the risk allele conveyed opposite effects for CP in our dataset, whereas this was only true for 2 of 9 in SNPs previously associated with CP. Although some of these findings could be a result of unclear effect allele reporting, there is evidence of opposite effects for the same allele in CL versus CP has been previously demonstrated near *IRF6*(13) (rs72741048, in Table 3), indicating additional investigation of these SNPs in the context of CL/P versus CP is warranted.

Our results support the hypothesis there are subtype-specific risks for CP, although this study has limitations. First, due to sample size we chose to evaluate clefts affecting both the hard and soft palate in both subtype groups, rather than as three separate groups. Given both structures are affected in these cases, it is likely they share risks for hard or soft palate clefts as suggested by our distinct findings between analyses. A lack of genome-wide significant signal in the ACP group could be due to our sample being underpowered to identify common variants of modest effect, or may result from SNPs of opposite effects negating signal when all phenotypes are combined. Alternatively, this may support a more prominent role for rare as opposed to common variants in the pathogenesis of CP in general, or there may be environmental effects not captured by our study. We also failed to replicate some well-established risk loci, such as *GRHL3* or *CTNNA2*, although this is likely explained by our study population. These risk variants occur in ∼3% of Europeans and ∼2% of Africans, respectively. In individuals of Asian ancestry, the MAF for rs41268753 in *GRHL3* is <1% (SAS) and <0.02% (EAS), and the MAF for both reported variants near *CTNNA2* (rs113691307 and rs80004662) is ∼4% (SAS) and <0.04% (EAS); therefore, our cohort is unlikely to harbor these variants at a rate detectable above our filter for common variants at MAF >3%. Despite these limitations, this is the first study to evaluate CP risks in the context of its subtypes and our findings show there are broad factors affecting the risk for cleft palate in general, as well as variants influencing the risk of specific CP subtypes.

## Methods

### Study population and phenotyping

The study population comes from multiple domestic and international sites where recruitment and phenotypic assessment occurred following institutional review board (IRB) approval for each local recruitment site and the coordinating center (University of Iowa, University of Pittsburgh, and Emory University). We assembled a collection of 435 case-parent trios ascertained on proband affection status (e.g., cleft palate) (Supplemental Table 1). The majority of trios (96%) consist of affected CP probands with unaffected parents. Although probands/trios were not excluded based on additional clinical features consistent with a syndromic diagnosis, only 45 trios were classified as possibly or probably syndromic based on a reported presence of additional major or minor clinical features. Trios represent all major ancestry groups affected by CP including those with European ancestry (recruited from Spain, Turkey, Hungary, United States), as well as understudied populations from Latin America (Puerto Rico, Argentina), Asia (China, Singapore, Taiwan, the Philippines), and Africa (Nigeria, Ghana). All probands and parents were assessed for the presence of a CP with ∼2/3 of the assembled samples undergoing additional phenotyping to assess the location and severity of the CP. Here we designate these probands as having a cleft of the hard and soft palate (n=82), cleft of the hard palate only (n=43), cleft of the soft palate only (n=152), and submucous cleft palate (n=25). For the purposes of analysis, submucous cleft palate was grouped with cleft soft palate.

### Sample preparation and whole genome sequencing

Whole genome sequencing was performed at the Center for Inherited Disease Research (CIDR) at Johns Hopkins University (Baltimore, MD). Prior to sequencing, samples were tested for adequate quantity and quality of genomic DNA using a Fragment Analyzer system and were processed with an Illumina InfiniumQCArray-24v1-0 array to confirm gender, relatedness, and known duplicates. For each sample, 500-750ng of genomic DNA was sheared to 400-600bp fragments, then processed with the Kapa Hyper Prep kit for End-Repair, A-Tailing, and Ligation of IDT (Integrated DNA Technologies) unique dual-indexed adapters according to the Kapa protocol to create a final PCR-free library.

A NovaSeq 6000 platform using 150bp paired-end runs was used for sequencing followed by base calling through the Illumina Real Time Analysis software (version 3.4.4). Files were demultiplexed from binary format (BCL) to individual fastq files with Illumina Isas bcl2fastq (version 1.37.1) and aligned to the human hg38 reference sequence (https://www.ncbi.nlm.nih.gov/assembly/GCF_000001405.39). The DRAGEN Germline v3.7.5 pipeline on the Illumina BaseSpace Sequence Hub platform was used for alignment, variant calling, and quality control, which produced single sample VCF files. The DRAGEN contamination detection tool was used to check for any cross-human sample contamination. Genotype concordance with existing array-based genotypes was performed using CIDRSeqSuite (version 7.5.0), and genotype concordance checks amongst replicate samples was performed in Picard GenotypeConcordance (Picard 2019). Following data quality steps and confirmation of adequate coverage (at least 80% of the genome at 20X or autosomal coverage at 30X), joint variant calling was performed, generating a multi-sample VCF file.

### Quality control

Variants aligning outside of standard chromosomes (1-22, X, Y), those with a filter flag and variants with a minor allele count (MAC) of <1 were removed. Genotypes with a quality score of <20 or a read depth of <10 were set to missing, and sites with missingness values of >10% were subsequently filtered out. Sample-level quality control metrics included transition/transversion (Ts/Tv) ratio, silent/replacement rate, and heterozygous/homozygous ratio; outlier samples with values outside of 3 standard deviations from the cohort mean were discarded. Samples with high missing data (>5% missing) were removed.

### Principal component analysis

A principal component (PC) analysis was performed for probands only using PLINK (v2.0) to determine genetic ancestry within our cohort. Variants were excluded if the minor allele frequency (MAF) <15% or if any site was missing a genotype (missingness >0%). We performed linkage disequilibrium (LD) pruning for R^2^ >0.1 prior to analysis. Ultimately, there were 67,584 variants for which we generated 15 PCs. After visualization of PC plots, PCs 1-3 were used to group by ancestry (Supplemental Figure 4, 5A).

### Statistical analysis

We performed transmission disequilibrium tests (TDT) to statistically analyze common variant associations with CP. The TDT, originally described by Spielman *et al*(34) tests for the rate of transmission of the minor allele to an affected proband using a McNemar’s test (i.e., a modified Chi-squared test for paired data). Because it tests transmission rather than allele frequency, it is robust to population stratification; however, it is only informative for sites at which parents are heterozygous for a variant. As differences in heterozygosity between populations may mask signals, we also performed a Chi-square test of homogeneity on our three CP groups to verify there were no significant differences in population makeup. Following filtering steps described above, the multisample VCF was imported into PLINK (version 1.90b53). Trios for which all individuals had a genotype missingness rate <5% and a Mendelian error rate <2% were included. Variants were included if they met the following criteria: MAF of >=3%, Mendelian error rate <0.1%, Hardy-Weinberg exact test (HWE) p-value of >1×10^−6^, and missingness rate of <5%. Results were considered genome-wide significant at p<5×10^−8^, and suggestive of significance at p<5×10^−6^. We report our odds ratio (OR) in reference to the alternate allele, and list the effect allele as that which increases risk for CP. Following TDT output, we applied FINEMAP (18) to our genome-wide significant locus. Briefly, FINEMAP uses a stochastic shotgun search to calculate posterior probability of SNP association with disease based on effect size (for which we used the natural log of the OR), MAF, and an LD matrix (generated in PLINK). We ran FINEMAP on SNPs within 1Mb in either direction of the lead SNP using default settings.

### DECIPHER variants

We queried the DECIPHER database (22) for copy number variants (CNVs) affecting *ANGPTL2* for individuals with phenotypes related to both palate and limb abnormalities. Terms included for palatal phenotypes were: cleft palate, high palate, narrow palate, narrow mouth, and micrognathia. Terms included for limb phenotypes were: 2-3 toe syndactyly, arachnodactyly, camptodactyly, long toe, abnormality of finger, tapered finger, upper limb undergrowth, and short foot. We then compared the rate of these phenotypes in individuals with CNVs to the general population based on the EUROCAT prevalence data using a two tailed Fisher’s exact test.

### Animal studies and gene expression assays

Animal studies were conducted in strict accordance with recommendations in the Guide for the Care and Use of Laboratory Animals of the National Institutes of Health. The protocol was approved by the University of Wisconsin School of Veterinary Medicine Institutional Animal Care and Use Committee (protocol number 13-081.0). C57BL/6J mice were purchased from The Jackson Laboratory and housed in rooms maintained at 22 ± 2 °C and 30–70% humidity on a 12 hour dark cycle. Mice were fed Irradiated Soy Protein-Free Extruded Rodent Diet (Catalog No. 2920x; Envigo Teklad Global) until day of plug, when dams received Irradiated Teklad Global 19% Protein Extruded Rodent Diet (Catalog No. 2919; Envigo Teklad Global). For timed matings, one or two nulliparous female mice were placed with a single male mouse for 1-2 hours and then examined for copulation plugs. The beginning of the mating period was designated as gestational day (GD)0, and pregnancy was confirmed by assessing weight gain between GD7 and GD10, as previously described (35). Dams were euthanized by carbon dioxide inhalation followed by cervical dislocation between GD10-14.5 ± 1 hour for embryo collection. One cohort of embryos collected for *in situ hybridization* assays were dissected in PBS and fixed in 4% paraformaldehyde for 18 h. Embryos subsequently underwent graded dehydration (1:3, 1:1, 3:1 v/v) into 100% methanol and were stored at -20°C indefinitely. Riboprobes were synthesized with gene-specific primers (Supplemental Table 3), and *in situ hybridization* was performed as previously described (36, 37). Embryos were subsequently embedded in 4% agarose gel and cut in sections (130 μM for head, 60 μM for limb) using a vibrating microtome. Images were captured using a MicroPublisher 5.0 camera (QImaging) mounted on an Olympus SZX-10 stereomicroscope. Another cohort of embryos was generated for quantitative gene expression analysis. Embryos were collected and microdissected in PBS, and enzymatic separation and isolation of the mesenchyme from maxillary process (GD10-12) or palatal shelf tissue (GD13-14) was performed as previously described (38, 39). RNA was isolated using the Qiagen RNeasy Mini Kit with on-column DNase I digestion according to the manufacturer recommendations. cDNA was synthesized from 100 ng of total RNA using the GoScript reverse transcription reaction kits (Promega). Singleplex quantitative real-time polymerase chain reaction (RT-qPCR) was performed using SSoFast EvaGreen Supermix (Bio-Rad) on a Bio-Rad CFX96 real-time PCR detection system (Bio-Rad Laboratories). RT-qPCR primers were designed using PrimerQuest (IDT), and sequences are listed in Supplemental Table 4. Target gene specificity was confirmed using National Center for Biotechnology Information Primer Basic Local Alignment Search Tool (NCBI Primer-BLAST). *Gapdh* was used as the housekeeping gene, and analyses were conducted with the 2^-ΔΔCt^ method.

### Replication of previously published SNPs

We searched for previously published SNPs associated with OFCs using the NHGRI-EBI GWAS catalog(23), which reports any SNPs with p-values less than 1×10^−5^. We initially identified 202 SNPs from GWAS data; however, after filtering for duplicates (i.e., SNPs reported in multiple studies) there were 166 SNPs of interest. When reporting the p-value for duplicated SNPs, we chose the most significant value. We then evaluated these variants for association with CP or CP subtypes in our current dataset and found 139 SNPs with data in at least one analysis. When comparing the two datasets, we reported the effect allele as the allele with increased OR as found in our current study.

## Data Availability

Sequence and phenotype data is available from the Database of Genotypes and Phenotypes (dbGaP) under study accession phs002220.v1.p1.

## Acknowledgements

We are incredibly grateful to participating families and colleagues who have made this research possible. Sequencing services were provided by the Center for Inherited Disease Research (CIDR). CIDR is fully funded through a federal contract from the National Institutes of Health to The Johns Hopkins University, contract number HHSN268201700006I. Patient recruitment, assembly of phenotypic information, sequencing services, and data analysis were supported by National Institutes of Health (NIH) grants: X01-HG010835 (EL), R01-DE016148 (MM, SW), R01-DE030342 (EL), R01-DE011931 (JH), R01-DE028300 (AB), R01-DE014581 (TB), R37-DE008559 (JM), R00-DE024571 (CB), S21-MD001830 (CB) U54-GM133807 (CB), T32-GM008490 (KR), F31-DE032588 (KR), R56-DE030917 (RL), F32-DE032260 (SWC). Some of this work was supported through cooperative agreements under PA 96043 from the Centers for Disease Control and Prevention to the Centers for Birth Defects Research and Prevention participating in the NBDPS. The findings and conclusions in this report are those of the authors and do not necessarily represent the official position of the Centers for Disease Control and Prevention or the California Department of Public Health. This article was prepared while Trenell J. Mosley was employed at Emory University. The opinions expressed in this article are the author’s own and do not reflect the view of the National Institutes of Health, the Department of Health and Human Services, or the United States government. This study makes use of data generated by the DECIPHER community. A full list of centers who contributed to the generation of the data is available from https://deciphergenomics.org/about/stats and via email from. Funding for the DECIPHER project was provided by Wellcome [grant number WT223718/Z/21/Z].

## References

1. Mossey PA, Modell B. Epidemiology of oral clefts 2012: an international perspective. Front Oral Biol. 2012;16:1–18.

2. Mai CT, Isenburg JL, Canfield MA, Meyer RE, Correa A, Alverson CJ, et al. National population-based estimates for major birth defects, 2010-2014. Birth Defects Res. 2019;111(18):1420–35.

3. Burg ML, Chai Y, Yao CA, Magee W, 3rd, Figueiredo JC. Epidemiology, Etiology, and Treatment of Isolated Cleft Palate. Frontiers in physiology. 2016;7:67-.

4. Berk NW, Marazita ML. Cleft Lip and Palate: From Origin to Treatment. Wyszynski DF, editor. Oxford: Oxford University Press; 2002.

5. Sivertsen A, Wilcox AJ, Skjaerven R, Vindenes HA, Abyholm F, Harville E, et al. Familial risk of oral clefts by morphological type and severity: population based cohort study of first degree relatives. BMJ (Clinical research ed). 2008;336(7641):432–4.

6. Grosen D, Chevrier C, Skytthe A, Bille C, Mølsted K, Sivertsen A, et al. A cohort study of recurrence patterns among more than 54,000 relatives of oral cleft cases in Denmark: support for the multifactorial threshold model of inheritance. Journal of medical genetics. 2010;47(3):162–8.

7. Grosen D, Bille C, Petersen I, Skytthe A, Hjelmborg J, Pedersen JK, et al. Risk of oral clefts in twins. Epidemiology. 2011;22(3):313–9.

8. Leslie EJ, Marazita ML. Genetics of cleft lip and cleft palate. Am J Med Genet C Semin Med Genet. 2013;163C(4):246–58.

9. Dixon MJ, Marazita ML, Beaty TH, Murray JC. Cleft lip and palate: understanding genetic and environmental influences. Nat Rev Genet. 2011;12(3):167–78.

10. Leslie EJ, Liu H, Carlson JC, Shaffer JR, Feingold E, Wehby G, et al. A Genome-wide Association Study of Nonsyndromic Cleft Palate Identifies an Etiologic Missense Variant in GRHL3. Am J Hum Genet. 2016;98(4):744–54.

11. Leslie EJ, Carlson JC, Shaffer JR, Butali A, Buxo CJ, Castilla EE, et al. Genome-wide meta-analyses of nonsyndromic orofacial clefts identify novel associations between FOXE1 and all orofacial clefts, and TP63 and cleft lip with or without cleft palate. Hum Genet. 2017;136(3):275–86.

12. Yu Y, Zuo X, He M, Gao J, Fu Y, Qin C, et al. Genome-wide analyses of non-syndromic cleft lip with palate identify 14 novel loci and genetic heterogeneity. Nat Commun. 2017;8:14364.

13. Huang L, Jia Z, Shi Y, Du Q, Shi J, Wang Z, et al. Genetic factors define CPO and CLO subtypes of nonsyndromicorofacial cleft. PLoS Genet. 2019;15(10):e1008357.

14. Butali A, Mossey PA, Adeyemo WL, Eshete MA, Gowans LJJ, Busch TD, et al. Genomic analyses in African populations identify novel risk loci for cleft palate. Hum Mol Genet. 2019;28(6):1038–51.

15. He M, Zuo X, Liu H, Wang W, Zhang Y, Fu Y, et al. Genome-wide Analyses Identify a Novel Risk Locus for Nonsyndromic Cleft Palate. J Dent Res. 2020;99(13):1461–8.

16. Rahimov F, Nieminen P, Kumari P, Juuri E, Nikopensius T, Karjalainen J, et al. High incidence and regional distribution of cleft palate in Finns are associated with a functional variant in an IRF6 enhancer. 2021.

17. Smith TM, Lozanoff S, Iyyanar PP, Nazarali AJ. Molecular signaling along the anterior-posterior axis of early palate development. Front Physiol. 2012;3:488.

18. Benner C, Spencer CCA, Havulinna AS, Salomaa V, Ripatti S, Pirinen M. FINEMAP: efficient variable selection using summary data from genome-wide association studies. Bioinformatics. 2016;32(10):1493–501.

19. Wilderman A, VanOudenhove J, Kron J, Noonan JP, Cotney J. High-Resolution Epigenomic Atlas of Human Embryonic Craniofacial Development. Cell Rep. 2018;23(23):1581–97.

20. Bush JO, Jiang R. Palatogenesis: morphogenetic and molecular mechanisms of secondary palate development. Development. 2012;139(139):231–43.

21. Rubin L, Saunders JW, Jr. Ectodermal-mesodermal interactions in the growth of limb buds in the chick embryo: constancy and temporal limits of the ectodermal induction. Dev Biol. 1972;28(28):94–112.

22. Firth HV, Richards SM, Bevan AP, Clayton S, Corpas M, Rajan D, et al. DECIPHER: Database of Chromosomal Imbalance and Phenotype in Humans Using Ensembl Resources. Am J Hum Genet. 2009;84(4):524–33.

23. Buniello A, MacArthur JAL, Cerezo M, Harris LW, Hayhurst J, Malangone C, et al. The NHGRI-EBI GWAS Catalog of published genome-wide association studies, targeted arrays and summary statistics 2019. Nucleic Acids Res. 2019;47(D1):D1005–D12.

24. Kent WJ, Sugnet CW, Furey TS, Roskin KM, Pringle TH, Zahler AM, et al. The human genome browser at UCSC. Genome Res. 2002;12(6):996–1006.

25. Takano A, Fukuda T, Shinjo T, Iwashita M, Matsuzaki E, Yamamichi K, et al. Angiopoietin-like protein 2 is a positive regulator of osteoblast differentiation. Metabolism -Clinical and Experimental. 2017;69:157–70.

26. Han X, Feng J, Guo T, Loh YE, Yuan Y, Ho TV, et al. Runx2-Twist1 interaction coordinates cranial neural crest guidance of soft palate myogenesis. Elife. 2021;10.

27. Zhang B, Chang J, Fu M, Huang J, Kashyap R, Salavaggione E, et al. Dosage effects of cohesin regulatory factor PDS5 on mammalian development: implications for cohesinopathies. PLoS One. 2009;4(5):e5232.

28. Kelly TJ, Suzuki HI, Zamudio JR, Suzuki M, Sharp PA. Sequestration of microRNA-mediated target repression by the Ago2-associated RNA-binding protein FAM120A. RNA. 2019;25(10):1291–7.

29. Tanaka M, Sasaki K, Kamata R, Hoshino Y, Yanagihara K, Sakai R. A novel RNA-binding protein, Ossa/C9orf10, regulates activity of Src kinases to protect cells from oxidative stress–induced apoptosis. Mol Cell Biol. 2009;29(2):402–13.

30. Ferguson MW, Sharpe PM, Thomas BL, Beck F. Differential expression of insulin-like growth factors I and II (IGF I and II), mRNA, peptide and binding protein 1 during mouse palate development: comparison with TGF beta peptide distribution. J Anat. 1992;181 (Pt 2):219–38.

31. Eggenschwiler J, Ludwig T, Fisher P, Leighton PA, Tilghman SM, Efstratiadis A. Mouse mutant embryos overexpressing IGF-II exhibit phenotypic features of the Beckwith-Wiedemann and Simpson-Golabi-Behmel syndromes. Genes Dev. 1997;11(23):3128–42.

32. Hoornaert KP, Vereecke I, Dewinter C, Rosenberg T, Beemer FA, Leroy JG, et al. Stickler syndrome caused by COL2A1 mutations: genotype-phenotype correlation in a series of 100 patients. Eur J Hum Genet. 2010;18(8):872–80.

33. Camargo M, Rivera D, Moreno L, Lidral AC, Harper U, Jones M, et al. GWAS reveals new recessive loci associated with non-syndromic facial clefting. Eur J Med Genet. 2012;55(10):510–4.

34. Spielman RS, McGinnis RE, Ewens WJ. Transmission test for linkage disequilibrium: the insulin gene region and insulin-dependent diabetes mellitus (IDDM). Am J Hum Genet. 1993;52(3):506–16.

35. Heyne GW, Plisch EH, Melberg CG, Sandgren EP, Peter JA, Lipinski RJ. A Simple and Reliable Method for Early Pregnancy Detection in Inbred Mice. J Am Assoc Lab Anim Sci. 2015;54(54):368–71.

36. Abler LL, Mehta V, Keil KP, Joshi PS, Flucus CL, Hardin HA, et al. A high throughput in situ hybridization method to characterize mRNA expression patterns in the fetal mouse lower urogenital tract. J Vis Exp. 2011(54).

37. Everson JL, Sun MR, Fink DM, Heyne GW, Melberg CG, Nelson KF, et al. Developmental Toxicity Assessment of Piperonyl Butoxide Exposure Targeting Sonic Hedgehog Signaling and Forebrain and Face Morphogenesis in the Mouse: An in Vitro and in Vivo Study. Environ Health Perspect. 2019;127(127):107006.

38. Everson JL, Fink DM, Yoon JW, Leslie EJ, Kietzman HW, Ansen-Wilson LJ, et al. Sonic hedgehog regulation of Foxf2 promotes cranial neural crest mesenchyme proliferation and is disrupted in cleft lip morphogenesis. Development. 2017;144(144):2082–91.

39. Li H, Williams T. Separation of mouse embryonic facial ectoderm and mesenchyme. J Vis Exp. 2013(74).

