## Supplemental Figure for "Trio-based GWAS identifies novel associations and subtype-specific risk factors for cleft palate"

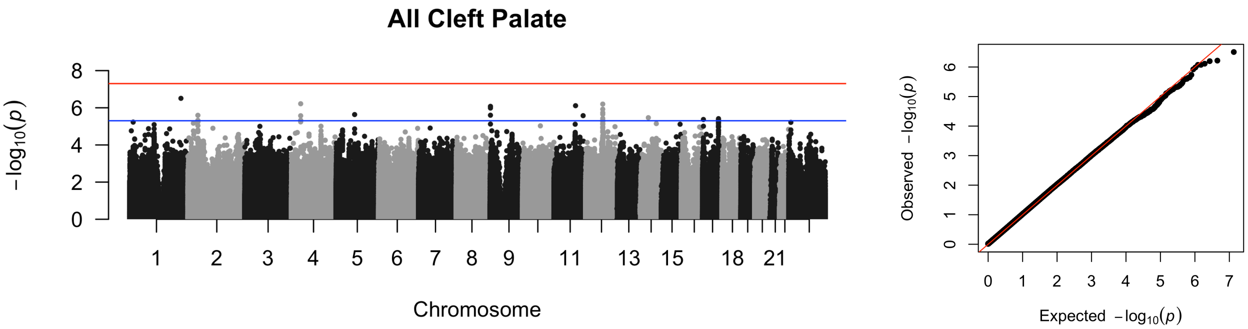


Supplemental Figure 1: Manhattan plot (left) and qqplot (right) for the ACP analysis. The blue line represents the suggestive threshold ($5x10^{-6}$) and the red line represents genome-wide significance ($5x10^{-6}$).

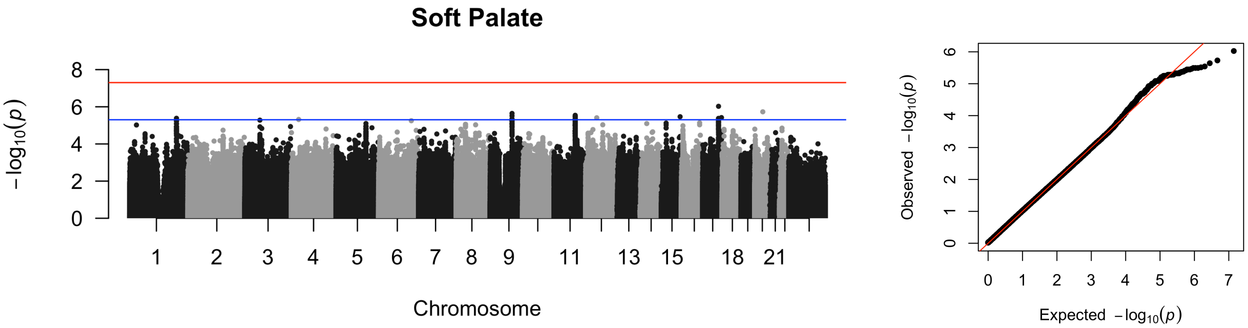


Supplemental Figure 2: Manhattan plot (left) and qqplot (right) for the CSP analysis. The blue line represents the suggestive threshold ($5x10^{-6}$) and the red line represents genome-wide significance ($5x10^{-6}$).

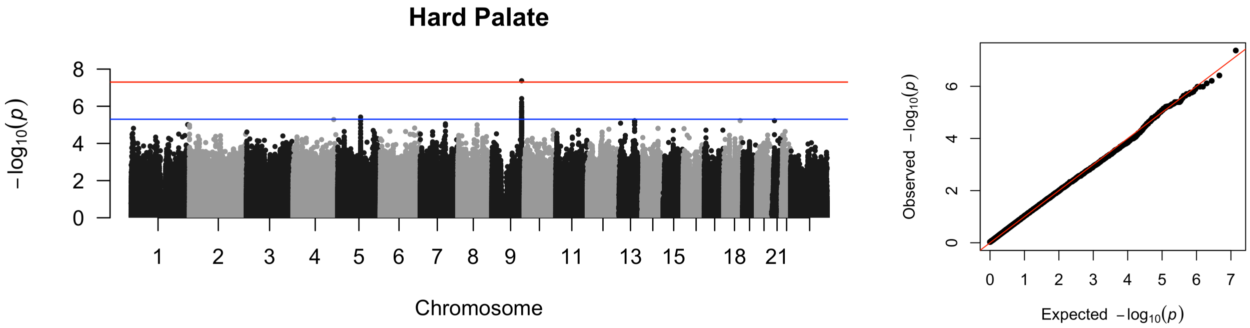


Supplemental Figure 3: Manhattan plot (left) and qqplot (right) for the CHP analysis. The blue line represents the suggestive threshold ($5x10^{-6}$) and the red line represents genome-wide significance ($5x10^{-6}$).

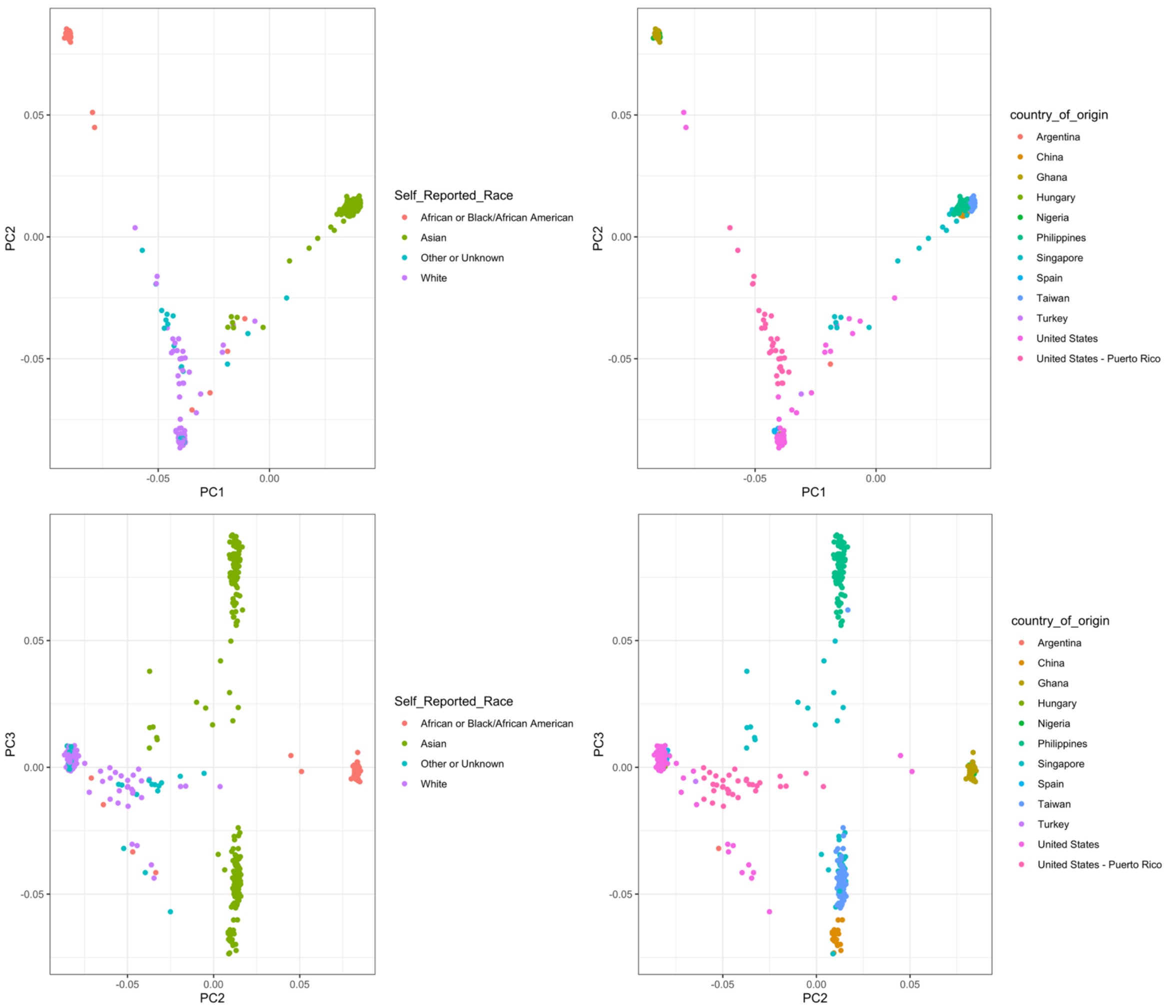


Supplemental Figure 4: Principal components 1 vs 2 (top) and 2 vs 3 (bottom) demonstrating group separation by self-reported race and by country of origin.

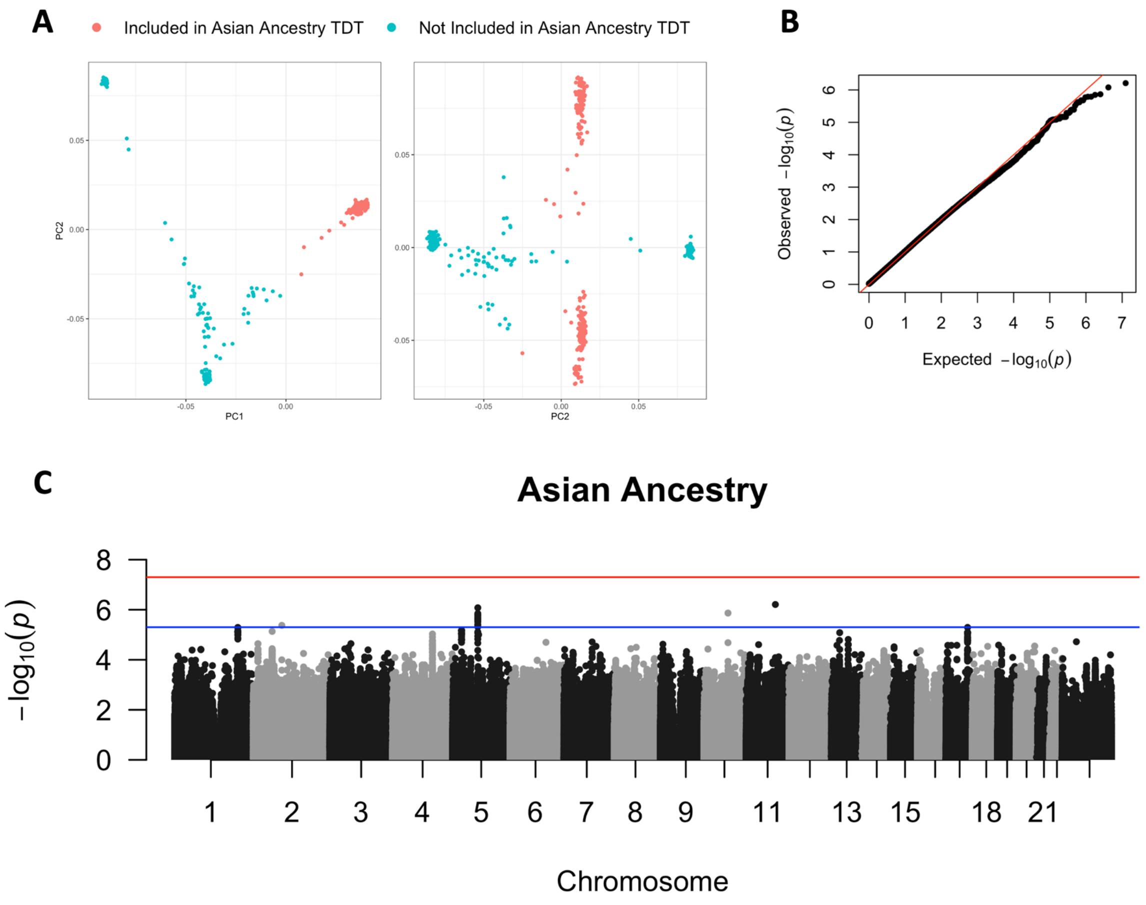


Supplemental Figure 5: A) Asian ancestry as determined by PC grouping – pink dots represent probands of Asian descent (n=262). B) qqplot for Asian ancestry-specific TDT. C) Manhattan plot for proband analysis. The blue line represents the suggestive threshold ($5x10^{-6}$) and the red [illegible]
